# Modelling information–dependent social behaviors in response to lockdowns: the case of COVID–19 epidemic in Italy

**DOI:** 10.1101/2020.05.20.20107573

**Authors:** Bruno Buonomo, Rossella Della Marca

## Abstract

The COVID–19 pandemic started in January 2020 has not only threatened world public health, but severely impacted almost every facet of lives including behavioral and psychological aspects. In this paper we focus on the ‘human element’ and propose a mathematical model to investigate the effects on the COVID–19 epidemic of social behavioral changes in response to lockdowns. We consider a SEIR-like epidemic model where that contact and quarantine rates are assumed to depend on the available information and rumors about the disease status in the community. The model is applied to the case of COVID–19 epidemic in Italy. We consider the period that stretches between Bebruary 24, 2020 when the first bulletin by the Italian Civil Brotection was reported and May 18, 2020 when the lockdown restrictions have been mostly removed. The role played by the information–related parameters is determined by evaluating how they affect suitable outbreak–severity indicators. We estimated that citizens compliance with mitigation measures played a decisive role in curbing the epidemic curve by preventing a duplication of deaths and about 46% more contagions.

**Subject class:** 92D30, 34C60

## 1 Introduction

In December 2019, the Municipal Health Commission of Wuhan, China, reported to the World Health Organization a cluster of viral pneumonia of unknown aetiology in Wuhan City, Hubei province. On January 9, 2020, the China CDC reported that the respiratory disease, later named COVID–19, was caused by the novel coronavirus SA RS CoY 2 [52]. The outbreak of COVID–19 rapidly expanded from Hubei province to the rest of China and then to other countries. Finally, it developed in a devastating pandemics affecting almost all the countries of the world [10]. As of May 18, 2020 a total of 4,7 million of cases of COVID–19 and 315,131 related deaths have been reported worldwide [10].

In absence of treatment and vaccine, the mitigation strategy enforced by many countries during the COVID–19 pandemic have been based on social distancing. Each government enacted a series of restriction affecting billions of people including recommendation of restricted movements for some or all of their citizens, and localized or national lockdown with the partial or full closing-off of non-essential companies and manufacturing plants [5].

Italy has been the first European country affected by COVID–19. The country has been strongly hit by the epidemic which has triggered progressively stricter restrictions aimed at minimizing the spread of the coronavirus. The actions enacted by the Italian government began with reducing social interactions through quarantine and isolation till to reach the *full lockdown* [1,30]. On May 4, 2020, the *phase two* began, marking a gradual reopening of the economy and restriction easing for residents. One week later, shops also reopened and the restrictions on mobility were essentially eliminated, with the only obligation in many regions to use protective masks [32].

During the period that stretches between January 22, 2020 and May 18, 2020, Italy suffered 225,886 official COVID–19 cases and 32,007 deaths [33].

The scientific community has promptly reacted to the COVID–19 pandemic. Since the early stage of the pandemic a number of mathematical models and methods has been used. Among the main concerns raised were: predicting the evolution of the COVID–19 pandemic wave worldwide or in specific countries [12,25,42]; predicting epidemic peaks and ICU accesses [46]; assessing the effects of containment measures [12,14,24,25,35,42,45] and, more generally, assessing the impact on populations in terms of economics, societal needs, employment, health care, deaths toll, etc [20,36].

Among the mathematical approaches used, many authors relied on deterministic compartmental models. This approach has been revealed successful for reproducing epidemic curves in the past SARS–CoV outbreak in 2002-2003 [26] and has been employed also for COVID–19.

Specific studies focused on the case of epidemic in Italy: Gatto *et al*. [24] studied the transmission between a network of 107 Italian provinces by using a SEPIA model as core model. Their SEPIA model discriminates between infectious individuals depending on presence and severity of their symptoms. They examined the effects of the intervention measures in terms of number of averted cases and hospitalizations in the period February 22 - March 25, 2020. Giordano *et al*. [25] proposed an even more detailed model, named SIDARTHE, in which the distinction between diagnosed and non-diagnosed individuals plays an important role. They used the SIDARTHE model to predict the course of the epidemic and to show the need to use testing and contact tracing combined to social distancing measures.

The mitigation measures like social distancing, quarantine and self-isolation may be encouraged or mandated [42]. However, although the vast majority of people were following the rules, even in this last case there are many reports of people breaching restrictions [4,47]. Local authorities needed to continuously verifying compliance with mitigation measures through monitoring by health officials and police actions (checkpoints, use of drones, fine or jail threats, etc). This behavior might be related to costs that individuals affected by epidemic control measures pay in terms of health, including loss of social relationships, psychological pressure, increasing stress and health hazards resulting in a substantial damage to population well-being [6,20,53].

As far as we know, the mathematically oriented papers on COVID–19 nowadays available in the literature do not explicitly take into account of the fraction of individuals that change their social behaviors solely in response to social alarm. From a mathematical point of view, the change in social behaviors maybe described by employing the method of information–dependent models [40,54] which is based on the introduction of a suitable *information index M* (*t*) (see [40,54]). This method has been applied to vaccine-preventable childhood diseases [16,54] and is currently under development (see [8,37,59] for very recent contributions).

In this paper, the main goal is to assess the effects on the COVID–19 epidemic of human behavioral changes during the lockdowns. To this aim we build up an information-dependent SEIR-like model which is based on the key assumption that the choice to respect the lockdown restrictions, specifically the social distance and the quarantine, is partially determined on fully voluntary basis and depends on the available information and rumors concerning the spread of the COVID–19 disease in the community.

A second goal of this manuscript is to provide an application of the information index to a specific field-case, where the model is parametrized and the solutions compared with official data.

We focus on the case of COVID–19 epidemic in Italy during the period that begins on February 24, 2020, when the first bulletin by the Italian Civil Protection was reported [33], includes the partial and full lockdown restrictions, and ends on May 18, 2020 when the lockdown restrictions have been mostly removed. We stress the role played by circulating information by evaluating the absolute and relative variations of disease-severity indicators as functions of the information-related parameters.

The rest of the paper is organized as follows: in Section 2 we introduce the model balance equations and information-dependent processes. Two critical epidemiological thresholds are computed in Section 3. Section 4 is devoted to model parametrization for numerical simulations, that are then shown and discussed in Section 5. Conclusions and future perspective are given in Section 6.

## 2 Model formulation

### 2.1 State variables and balance equations

We assume that the total population is divided into seven disjoint *compartments*, susceptibles *S*, exposed *E*, pre-symptomatic *I_p_*, asymptomatic/mildly symptomatic *I_m_*, severely symptomatic (hospitalized) *I_s_*, quarantined *Q* and recovered *R*. Any individual of the population belongs to one (and only one) compartment.

The size of each compartment at time *t* represents a *state variable* of a mathematical model. The state variables and the processes included in the model are illustrated in the flow chart in Fig. 1. The model is given by the following system of nonlinear ordinary differential equations, where each (*balance equation*) rules the rate of change of a state variable.

**Figure 1:**
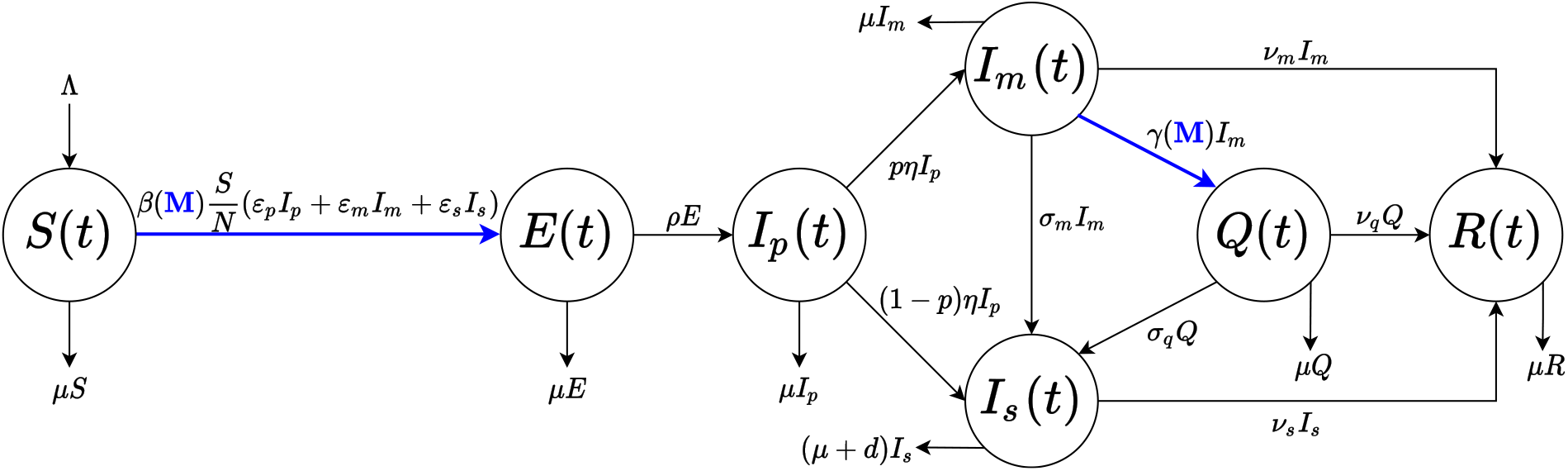
Flow chart for the COVID–19 model (1)–(3). The population is divided into seven disjoint compartments of individuals: susceptible *S*(*t*), exposed *E*(*t*), pre-symptomatic *I_p_*(*t*), asymptomatic/mildly symptomatic *I_m_*(*t*), severely symptomatic/hos pitalized *I_s_*(*t*), quarantin ed *Q*(*t*) and recove red *R*(*t*). Blue colour indicates the information–dependent processes in model (see (7)–(8)–(9), with *M*(*t*) ruled by (3)).

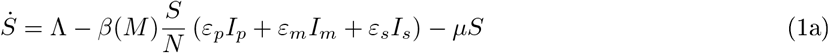

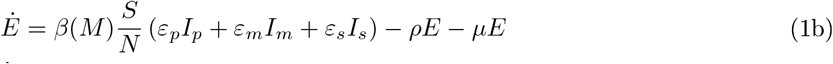

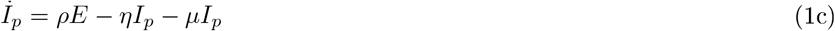

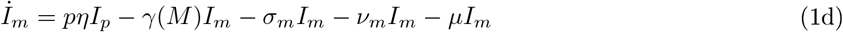

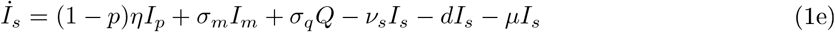

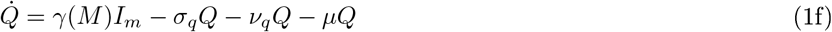

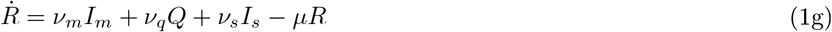

The model formulation is described in detail in the next subsections.

### 2.2 The role of information

As mentioned in the introduction, the mitigation strategy enforced by many countries during the COVID–19 pandemic has been based on social distancing and quarantine. Motivated by the discussion above, we assume that the final choice to adhere or not to adhere lockdown restrictions is partially determined on fully voluntary basis and depends on the available information and rumors concerning the spread of the disease in the community.

From a mathematical point of view, we describe the change in social behaviors by employing the method of information-dependent models [40,54]. The information is mathematically represented by an *information index M*(*t*) (see Appendix A for the general definition), which summarizes the information about the current and past values of the disease [8,15–17] and is given by the following distributed delay

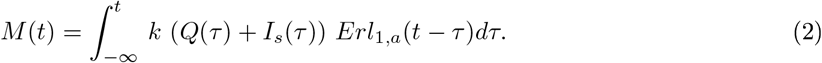

This formulation may be interpreted as follows: the first order Erlang distribution *Erl*_1_*,_a_*(*x*) represents an exponentially fading memory, where the parameter *a* is the inverse of the average time delay of the collected information on the status of the disease in the community (see Appendix A). On the other hand, we assume that people react in response to information and rumors regarding the daily number of quarantined and hospitalized individuals. The *information coverage k* is assumed to be positive and *k* ≤ 1, which mimics the evidence that COVID–19 official data could be under–reported in many cases [38,42]. With this choice, by applying the linear chain trick [39], we obtain the differential equation ruling the the *M*:

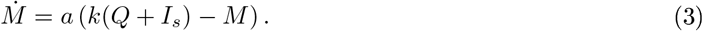

### 2.3 Formulation of the balance equations

In this section we derive in details each balance equation of model (1).

*Equation (1a): Susceptible individuals, S*(*t*)

Susceptibles are the individuals who are healthy but can contract the disease. Demography is incorporated in the model so that a net inflow rate 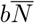 due to births is considered, where *b* is the birth rate and 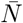 denotes the total population at the beginning of the epidemic.

We also consider an inflow term due to immigration, Λ_0_. Since global travel restrictions were implemented during the COVID–19 epidemic outbreak [50], we assume that Λ_0_ accounts only of repatriation of citizens to their countries of origin due to the COVID–19 pandemic [21]. In all airports, train stations, ports and land borders travellers’ health conditions were tested via thermal scanners. Although the effectiveness of such screening method is largely debated [43], for the sake of simplicity, we assume that the inflow enters only into the susceptibles compartment.

In summary, we assume that the total inflow rate Λ is given by:

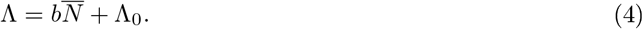

The susceptible population decreases by natural death, with death rate *μ* and following infection. It is believed that COVID–19 is primarily transmitted from symptomatic people (mildly or severely symptomatic). In particular, although severely symptomatic individuals are isolated from the general population by hospitalization, they are still able to infect hospitals and medical personnel [2,23] and, in turn, give rise to transmission from hospital to the community. The pre-symptomatic transmission (i.e. the transmission from infected people before they develop significant symptoms) is also relevant: specific studies revealed an estimate of 44% of secondary cases during the presymptomatic stage from index cases [27].

On the contrary, the asymptomatic transmission (i.e. the contagion from a person infected with COVID–19 who does not develop symptoms) seems to play a negligible role [55]. We also assume that quarantined individuals are fully isolated and therefore unable to transmit the disease.

The routes of transmission from COVID–19 patients as described above are included in the *Force of Infection* (FoI) function, i.e. the per capita rate at which susceptibles contract the infection:

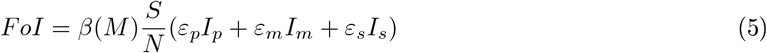

The transmission coefficients for these three classes of infectious individuals are information–dependent and given by *ε_p_β*(*M*), *ε_m_β*(*M*) and *ε_s_β*(*M*), respectively, with 0 ≤ *ε_p_, ε_m_*, *ε_s_* < 1.

The function *β*(*M*), which models how the information affects the transmission rate, is defined as follows: the baseline transmission rate *β*(·) is a piecewise continuous, differentiable and decreasing function of the information index *M*, with *β*(max(*M*)) > 0. We assume that

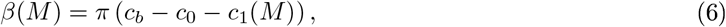

Where *π* is the probability of getting infected during a person–to–person contact and *c_b_* is the baseline contact rate. In (6) the reduction in social contacts is assumed to be the sum of a constant rate *c*_0_, which represent the individuals’ choice to self-isolate regardless of rumors and information about the status of the disease in the population, and an information-dependent rate *c*_1_(*M*), being *c*_1_(·) increasing with *M* and *c*_1_(0) = 0. In order to guarantee the positiveness, we assume *c_b_* > *c*_0_ + max(*c*_1_(*M*)). Following [15], we finally set

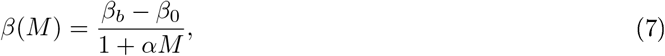

namely *πc_b_* = *β_b_*, *πc*_0_ = *β*_0_ and *πc*_1_(*M*) = *αM*(*β_b_* − *β*_0_)/(1 + *αM*), where *α* is a positive constant tuning the degree of voluntary change in contact patterns. For illustrative purposes, see Fig. 2, top panel.

**Figure 2:**
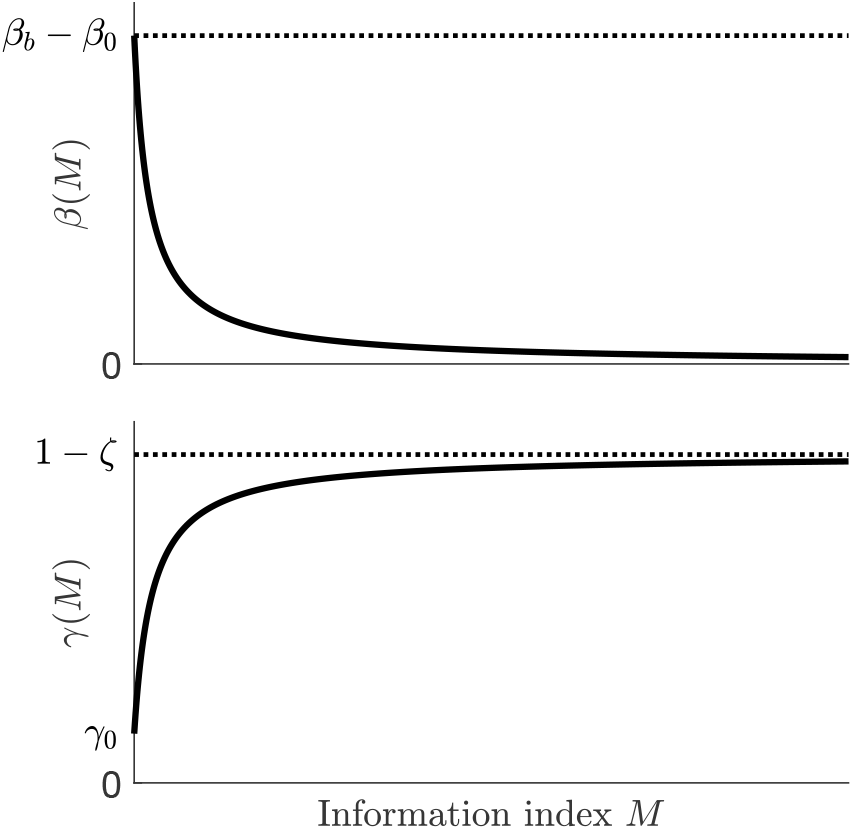
Representative shapes of the transmission rate (7) (top panel) and the quarantine rate (8) (bottom panel) as functions of the information index *M*.

#### *Equation* (*lb*)*: Exposed individuals, E*(*t*)

Exposed (or latent) individuals are COVID–19 infected but are not yet infectious, i.e. capable of transmitting the disease to others. Such individuals arise as the result of new infections of susceptible individuals. The population is diminished by development at the infectious stage (at rate *ρ*) and natural death.

#### *Equation* (*1c*)*: Pre–symptomatic individuals, I_p_*(*t*)

Pro-symptomatic individuals are infectious people that have not yet developed significant symptoms. Such individuals lie in a stage between the exposed and the expected symptomatic ones. They remain in this compartment, *I_p_*, during the post-latent incubation period and diminish due to natural death or progress to become asymptomatic or symptomatic infectious individuals (at a rate *η*).

#### *Equation* (*1d*)*: Asymptomatic/mildly symptomatic individuals, I_m_*(*t*)

This compartment includes both the asymptomatic individuals, that is infected individuals who do not develop symptoms, and mildly symptomatic individuals [24]. As mentioned above, the asymptomatic transmission seems to play a negligible role in COVID–19 transmission. However, asymptomatic individuals are infected people which results positive cases to screening (positive pharyngeal swabs) and therefore enter in the official data count of confirmed diagnosis. Members of this class come from pro symptomatic stage and get out due to quarantine (at an information-dependent rate γ(*M*)), worsening symptoms (at rate *σ_m_*) and recovery (at rate *ν_m_*).

#### *Equation* (*1e*)*: Severely symptomatic individuals* (*Hospitalized*)*, I_s_*(*t*)

Severely symptomatic individuals are isolated from the general population by hospitalization. They arise: *(i)* as consequence of the development of severe symptoms by mild illness (the infectious of the class *I_m_* or the quarantined *Q*); (*ii*) directly from the fraction 1 − *p* of pre-symptomatic individuals that rapidly develop in severe illness. This class diminishes by recovery (at rate *ν_s_*), natural death and disease-induced death (at rate *d*).

#### *Equation* (*1f*)*: Quarantined individuals, Q*(*t*)

Quarantined individuals *Q* are those who are separated from the general population. We assume that quarantined are asymptomatic/mildly asymptomatic individuals. This population is diminished by natural deaths, aggravation of symptoms (at rate *σ_q_*, so that they move to *I_s_*) and recovery (at a rate *ν_q_*). For simplicity, we assume that the quarantine is 100% effective, i.e. with no possibility of contagion. Quarantine may arise in two different ways. From one hand, individuals may be detected by health authorities and daily checked. Such health active surveillance ensures also that the quarantine is, in some extent, respected. On the other hand, a fraction of quarantined individuals choose self-isolation since they are confident in the government handling of crisis or just believe the public health messaging and act in accordance [3].

As mentioned in subsection 2.2, we assume that the final choice to respect or not respect the self-isolation depends on the awareness about the status of the disease in the community. Therefore, we define the information-dependent quarantine rate as follows. We assume that

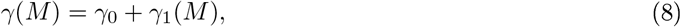

where the rate γ_0_ mimics the fraction of the asymptomatic/mildly symptomatic individuals *I_m_* that has been detected through screening tests and is ‘forced’ to home isolation. The rate γ_1_(*m*) represents the undetected fraction of individuals that adopt quarantine by voluntary choice as result of the influence of the circulating information *M*. The function γ_1_(·) is required to be a continuous, differentiable and increasing function w.r.t. *M*, with γ_1_(0) = 0. As in [8,16], we set

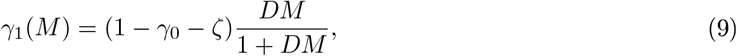

with *D >* 0 0 *<* ζ *<* 1 − γ_0_> potentially implying a roof of 1 − ζ in quarantine rate under circumstances of high perceived risk. A representative trend of γ(*m*) is displayed in Fig. 2, bottom panel.

*Equation* (*1g*)*: Recovered, individuals, R*(*t*)

After the infectious period, individuals from the compartments *I_m_*, *I_s_* mid *Q* recover at rates *ν_m_, ν_s_* and *ν_q_*, respectively. Natural death rate is also considered. We assume that individuals who recover from COVID–19 acquire long lasting immunity against COVID–19 although this is a currently debated question (as of 16 May, 2020) and there is still no evidence that COVID–19 antibodies may protect from re-infection [58].

## 3 The reproduction numbers

A frequently used indicator for measuring the potential spread of an infectious disease in a community is the *basic reproduction number*, 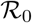, namely the average number of secondary cases produced by one primary infection over the course of infectious period in a fully susceptible population. If the system incorporates control strategies, then the corresponding quantity is named *control reproduction number* and usually denoted by 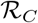 (obviously, 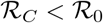).

The reproduction number can be calculated as the spectral radius of the *next generation* matrix *FV*^−1^, where *F* and *V* are defined as Jacobian matrices of the new infections appearance and the other rates of transfer, respectively, calculated for infected compartments at the disease-free equilibrium [13,51]. In the specific case, if *β*(*M*) *= β_b_* mid γ(*M*) *=* 0 in (1)–(3), namely when containment interventions are not enacted, we obtain the expression of 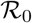; otherwise, the corresponding 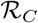 can be computed. Simple algebra yields

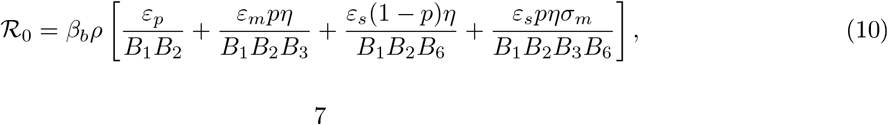

and

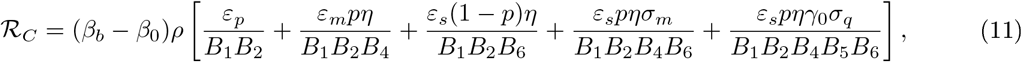

With

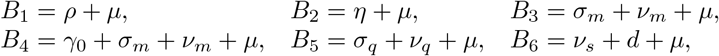

(for more details see Appendix B). The first two terms in the r.h.s of (11) describe the contributions of pre-symptomatic infectious and asymptomatic/mildly symptomatic infectious, respectively, to the production of new infections close to the disease-free equilibrium. The last three terms represent the contribution of infectious with severe symptoms, which could onset soon after the incubation phase or more gradually after a moderate symptomatic phase or even during the quarantine period. Note that the latter term is missing in the basic reproduction number (10), where the possibility for people to be quarantined is excluded. Note also that 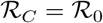 when *β*_0_= γ_0_ = 0.

## 4 Parametrization

Numerical simulations are performed in MATLAB environment [41] with the use of platform integrated functions. A detailed model parametrization is given in the next subsections.

### 4.1 Epidemiological parameters

The epidemiological parameters of the model as well as their baseline values are reported in Table 1. The most recent data by the Italian National Institute of Statistics [29] refer to January 1, 2019 and provide a country-level birth rate *b* = 7.2/1000 years^−1^ and a death rate *μ* = 10.7/1000 years^−1^, as well as a resident population of about

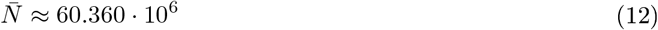

inhabitants. Fluctuations in a time window of just over a year are considered negligible.

**Table 1:** Epidemiological parameters values for model (1)–(3).

| Parameter | Description | Baseline value |
| --- | --- | --- |
| $\Lambda$ | Input term | $1.762 \cdot 10^3 \text{ days}^{-1}$ |
| $\mu$ | Death rate | $10.7/1000 \text{ years}^{-1}$ |
| $\mathcal{R}_0$ | Basic reproduction number | 3.49 |
| $\beta_b$ | Baseline transmission reduction | $2.25 \text{ days}^{-1}$ |
| $\beta_0$ | Rate of mandatory transmission reduction | $0 - 0.74\beta_b$ |
| $\varepsilon_p$ | Modification factor w.r.t. transmission from $I_p$ | 1 |
| $\varepsilon_m$ | Modification factor w.r.t. transmission from $I_m$ | 0.033 |
| $\varepsilon_s$ | Modification factor w.r.t. transmission from $I_s$ | 0.034 |
| $\rho$ | Latency rate | $1/5.25 \text{ days}^{-1}$ |
| $\eta$ | Post-latency rate | $1/1.25 \text{ days}^{-1}$ |
| $p$ | Fraction of pre-symptomatics developing no/mild symptoms | 0.92 |
| $\gamma_0$ | Rate of mandatory quarantine | $0.057 \text{ days}^{-1}$ |
| $\sigma_m$ | Rate at which asymptomatics/mildly symptomatics hospitalize | $0.044 \text{ days}^{-1}$ |
| $\sigma_q$ | Rate at which quarantined individuals hospitalize | $0.001 \text{ days}^{-1}$ |
| $d$ | Disease-induced death rate | $0.022 \text{ days}^{-1}$ |
| $\nu_m$ | Recovery rate for asymptomatic/mildly symptomatic individuals | $0.145 \text{ days}^{-1}$ |
| $\nu_q$ | Recovery rate for quarantined individuals | $0.035 \text{ days}^{-1}$ |
| $\nu_s$ | Recovery rate for heavily symptomatic/hospitalized individuals | $0.048 \text{ days}^{-1}$ |

The immigration inflow term Λ_0_ accounts for the repatriation of Italians abroad. On the basis of data communicated by the Italian Ministry of Foreign Affairs and International Cooperation [31], a reasonable value for Λ_0_ seems to be Λ_0_ = 4000/7 day^−1^, namely the average number of repatriated citizens is 4000 *per* week. From (4), we finally obtain Λ ≈ 1.762 · 10^3^ days^−1^.

Epidemiological data are based on the current estimates disseminated by national and international health organizations [19,28,34,42,56,57] or inferred by modelling studies [12,24,35]. More precisely, the median incubation period is estimated to be from 5–6 days, with a range from 1–14 days, and identification of the virus in respiratory tract specimens occurred 1–2 days before the onset of symptoms [19,56]. Hence, we set the latency (*ρ*) and pre-latency (*η*) rates to 1/5.25 days^−1^ and 1/1.25 days^−1^, respectively. From [24], the specific information-independent transmission rates for the pre-symptomatic (*ε_p_β_b_*), asymptomatic/mildly symptomatic (*ε_m_β_b_*) and severely symptomatic (*ε_s_β_b_*) cases are such that *ε_m_*/*ε_p_* = 0.033 and *ε_s_*/*ε_m_* = 1.03. They are in accordance with the observation of high viral load close to symptoms onset (suggesting that SARS-CoV-2 can be easily transmissible at an early stage of infection), and with the absence of reported significant difference in viral load in pre-symptomatic and symptomatic patients [19]. We set *β_b_* = 2.25 days^−1^, which, together with the other parameters, leads to the basic reproduction number 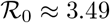, a value falling within the ranges estimated in [19,24,42,56].

As in [35], we consider that just 8% of infectious individuals show serious symptoms immediately after the incubation phase, yielding *p* = 0.92. Nonetheless, people with initial mild symptoms may become seriously ill and develop breathing difficulties, requiring hospitalization. It is estimated that about 1 in 5 people with COVID–19 show a worsening of symptoms [34] within 4–5 days from onset [28], giving *σ_m_* = 0.2/4.5 ≈ 0.044 days^−1^. Instead, the possibility that the aggravation happens during the quarantine period is assumed to be more rare: *σ_q_* = 0.001 days^−1^.

Governmental efforts in identifying and quarantining positive cases were implemented since the early stage of epidemics (at February 24, 94 quarantined people were already registered [33]), hence we consider the daily mandatory quarantine rate of asymptomatic/mildly symptomatic individuals (γ_0_) for the whole time horizon. From current available data, it seems hard to catch an uniform value for γ_0_ because it largely depends on the sampling effort, namely the number of specimen collections (swabs) from persons under investigation, that varies considerably across Italian regions and in the different phases of the outbreak [28,33]. Since our model does not account for such territorial peculiarities and in order to reduce the number of parameters to be estimated, we assume that γ_0_ = 1.3*σ_q_*, namely it is 30% higher than the daily rate at which members of the *I_m_* class hospitalize, yielding γ_0_ ≈ 0.057 days^−1^. Simulations with such a value provide a good approximation of the time-evolution of registered quarantined individuals at national level, as displayed in Fig. 4, second panel.

Following the approach adopted in [26] for a SARS-CoV epidemic model, we estimate the disease-induced death rate as

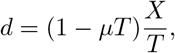

where *X* is the case fatality and *T* is the expected time from hospitalization until death. From [28], we approximate *X* = 13% and *T* = 6 days (it is 9 days for patients that were transferred to intensive care and 5 days for those were not), yielding *d* ≈ 0.022 days^−1^. Similarly, the recovery rates *ν_j_* with *j* ∈ {*m, q, s*} are estimated as

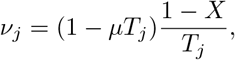

where *T_j_* is the expected time until recovery or expected time in quarantine/hospitalization. Preliminary data indicate that the virus can persist for up to eight days from the first detection in moderate cases and for longer periods in more severe cases [19], suggesting *T_m_* = 6 days is an appropriate value. As far as the time spent in hospitalization or quarantine, in the lack of exact data, we assume *T_s_ <T_q_*, because hospitalized individuals are likely to receive a partly effective, experimental treatment: mainly antibiotics, antivirals and corticosteroids [28]. Moreover, shortages in hospital beds and intensive care units (ICUs) lead to as prompt as possible discharge [22]. In particular, we set *T_s_* = 18 and *T_q_* = 25 days, by accounting also for prolonged quarantine time due to delays in test response (if any) and for WHO recommendations of an additional two weeks in home isolation even after symptoms resolve [57].

Crucially, we also estimate the initial exponential rate of case increase (say, *g*_0_), by computing the dominant eigenvalue of the system’s Jacobian matrix, evaluated at the disease-free equilibrium. It provides g_0_ ≈ 0.247 days^−1^, in accordance to that given in [24].

### 4.2 The lockdowns effect on the transmission

We explicitly reproduce in our simulations the effects of the progressive restrictions posed to human mobility and human to human contacts in Italy. Their detailed sequence may be summarized as follows. After the first officially confirmed case (the so called ‘patient one’) on February 21, 2020 in Lodi province, several suspected cases emerged in the south and southwest of Lombardy region. A ‘red zone’ encompassing 11 municipalities was instituted on February 22 and put on lockdown to contain the emerging threat. On March 8, the red zone was extended to the entire Lombardy and 14 more northern Italian provinces, while the rest of Italy implemented social distancing measures. A leak of a draft of this decree prompted a panic reaction with massive movement of people towards Italian regions, especially from the north to the south [49]. The next day, a decree evocatively entitled ‘I’m staying at home’ was signed: the lockdown was declared for the whole country with severe limitations to mobility and other progressively stricter restrictions.

Soon after, on March 11, 2020, the lockdown was extended to the entire country [30], with all commercial and retail businesses – except those providing essential services – closed down [30]. Finally, on March 22, 2020, the *phase one* of restrictions was completed when a *full* lockdown was imposed by closing all non essential companies and industrial plants [1]. On May 4, Italy entered the *phase two*, representing the starting point of a gradual relaxation of the restriction measures. One week later, shops also reopened and the restrictions on mobility were essentially eliminated, with the only obligation in many regions to use protective masks [32].

Because data early in an epidemic are inevitably incomplete and inaccurate, our approach has been to try to focus on what we believe to be the essentials in formulating a simple model. Keeping this in mind, we assume that the disease transmission rate incurs in just two step reductions (modelled by the reduction rate *β*_0_ in (7)), corresponding to

- March 12 (day 17), when the lockdown decree came into force along with the preceding restrictions, cumulatively resulting in a sharp decrease of SARS-CoV-2 transmission;
- March 23 (day 28), the starting date of the *full* lockdown that definitely impacted the disease incidence.

In the wake of [25,42], we account for a first step reduction by 64% (that is 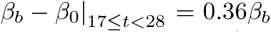), which drops the control reproduction number (3) close to 1 (see Fig. 3, dotted black and red lines). It is then strengthened by an additional 28% about, resulting in a global reduction by 74% 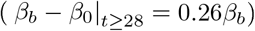 that definitely brings 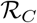 below 1 (see Fig. 3, dotted black and blue lines).

**Figure 3:**
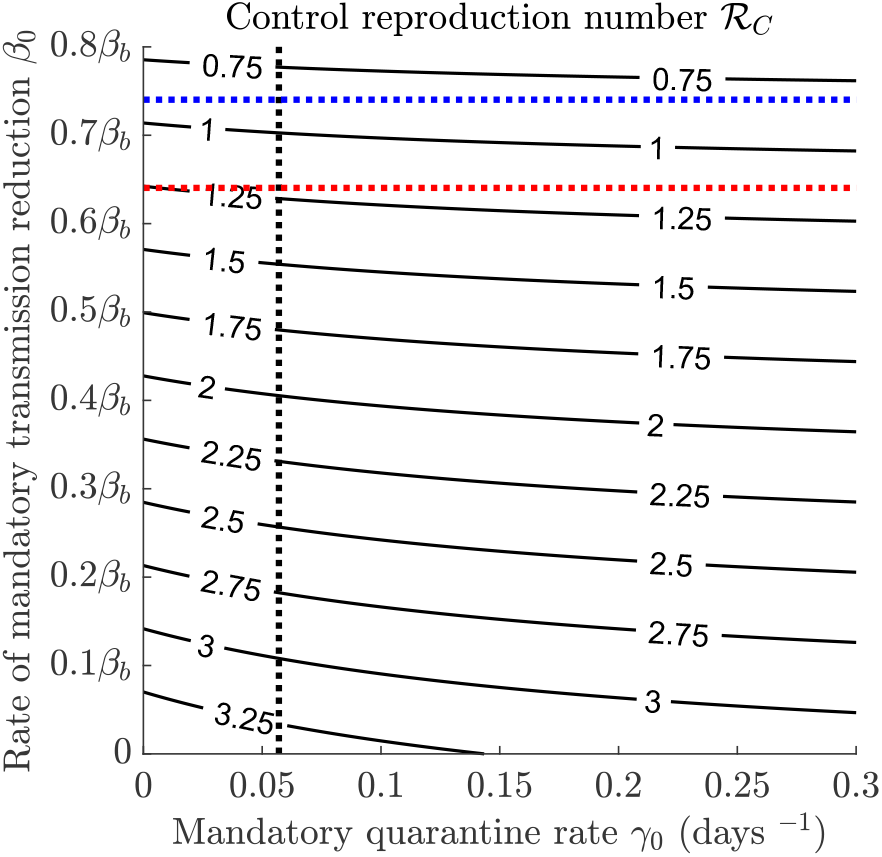
Contour plot of the control reproduction number (11) versus mandatory quarantine and transmission reduction rates. Intersection between dotted black and red (resp. blue) lines indicates the value after the first (resp. second) step reduction. Other parameters values are given in Table 1.

### 4.3 Information dependent parameters

The information-related parameter values are reported in Table 2 together with their baseline values. Following [8,16], we set ζ = 0.01 days^−1^ potentially implying an asymptotic quarantine rate of 0.99 days^−1^ if we could let *M* go to +∞. The positive constants *α* e *D* tune the information-dependent *reactivity*, respectively, of susceptible and infectious people in reducing mutual social contacts and of individuals with no/mild symptoms in self-isolating. In virtue of the order of magnitude of the information index *M* (ranging between 10 Mid 10^5^), we set *α* = 6 · 10^−7^ and *D* = 9 · 10^−6^, resulting in a receptive propensity to self-isolation for asymptomatics/mildly symptomatics and less evident degree of variability in contact rate, being the latter already impacted and constrained by government laws (as shown later in Fig. 7). Values range for the information coverage *k* and the average time delay of information 1/*a* are mainly guessed or taken from papers where the information index *M* is used [7,8,15,16]. The former may be seen as a ‘summary’ of two opposite phenomena: the disease under-reporting and the level of media coverage of the status of the disease, which tends to amplify the social alarm. It is assumed to range from a minimum of 0.2 (i.e. the public awareness is of 20%) to 1. The latter ranges from the hypothetical case of prompt communication (*a* = 1 days^−1^) to a delay of 60 days. We tune these two parameters within their values range in order to reproduce the curves that best fit with the number of hospitalized individuals (*I_s_*) and the cumulative deaths as released every day at 6 p.m. (UTC+1 h) since February 24, 2020 by the Italian Civil Protection Department and archived on GitHub [33].

**Table 2:** Information-dependent parameters values for model (1)–(3).

| Parameter | Description | Baseline value |
| --- | --- | --- |
| $\alpha$ | Degree of voluntary change in contact patterns | $6 \cdot 10^{-7}$ |
| $D$ | Reactivity factor of voluntary quarantine | $9 \cdot 10^{-6}$ |
| $\zeta$ | $1 - \zeta$ is the roof of overall quarantine rate | $0.01 \text{ days}^{-1}$ |
| $a$ | $1/a$ is the average information delay | $1/3 \text{ days}^{-1}$ |
| $k$ | Degree of information coverage | 0.8 |

We find a good approximation by setting *k* = 0.8 and *a* = 1/3 days^−1^, meaning a level of awareness about the daily number of quarantined and hospitalized of 80%, resulting from the balance between underestimates and media amplification and inevitably affected by rumors and misinformation spreading on the web (the so-called ‘infodemic’ [44]). Such awareness is not immediate, but information takes on average 3 days to be publicly disseminated, being the communication slowed by a series of articulated procedures: timing for swab tests results, notification of cases, reporting delays between surveillance and public health authorities, and so on.

Of course, parameters setting is influenced by the choice of curves to fit. Available data seem to provide an idea about the number of identified infectious people who have developed mild/moderate symptoms (the fraction that mandatorily stays in *Q*) or more serious symptoms (the hospitalized, *I_s_*) and the number of deaths, but much less about those asymptomatics or with very mild symptoms who are not always subjected to a screening test.

### 4.4 Initial conditions

In order to provide appropriate initial conditions, we consider the official national data at February 24, 2020 archived on [33]. In particular, we take the number of mandatorily quarantined individuals (at that time, they coincide with *Q* being the voluntary component negligible) and the hospitalized people (*I_s_*). Then, we simulate the temporal evolution of the epidemics prior to February 24, by imposing an initial condition of one exposed case Δ*t*_0_ days before in a population of 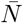 individuals, with 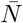 given in (12). We assume *β*_0_ = 0 and γ_0_ as in Table 1 (no social distance restrictions were initially implemented, but quarantine efforts were active since then) and disregard the effect of information on the human social behaviors in this phase (*α* = *D* = 0 in (1)–(3)). The length of temporal interval Δ*t*_0_ is tuned in order to reproduce the official values released for *Q* and *I_s_* at February 24 and provide estimations for the other state variables, as reported in Table 3. We obtain Δ*t*_0_ = 31.9, indicating that the virus circulated since the end of January, as predicted also in [11,24].

**Table 3:**
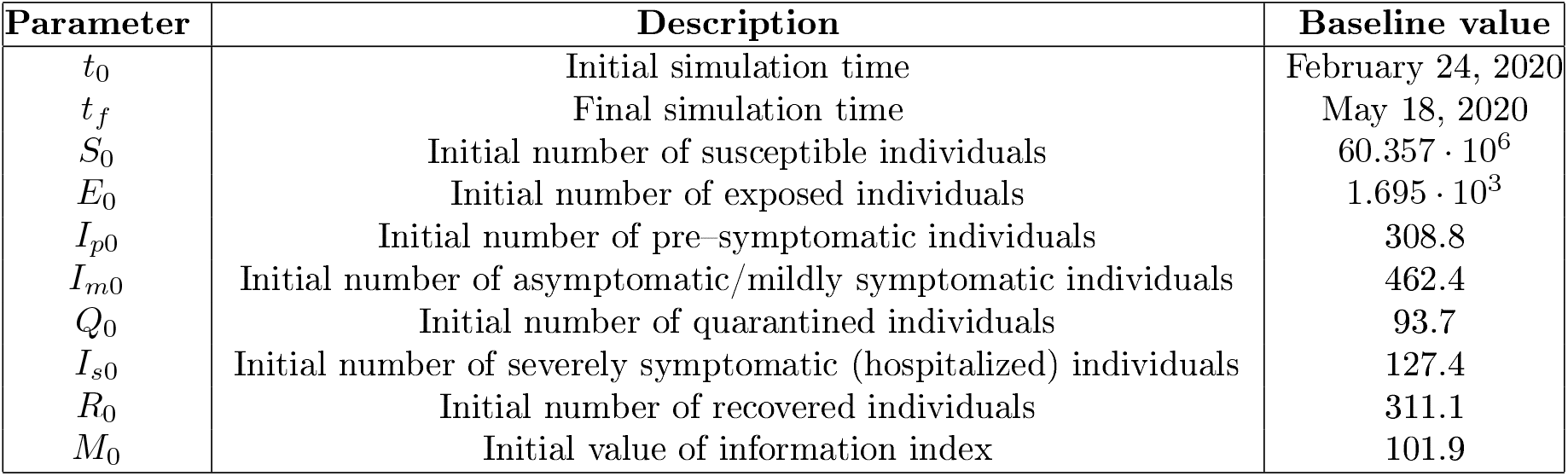
Temporal horizon and initial conditions for model (1)–(3).

## 5 Numerical results

Let us consider the time frame [*t*_0_,*t*], where *t*_0_ ≤ *t* ≤ *t_f_*. We consider two relevant quantities, the *cumulative incidence* CI(*t*), i.e. the total number of new cases in [*t*_0_,*t*], and the *cumulative deaths* CD(*t*), i.e. the disease-induced deaths in [*t*_0_,*t*].

For model (1)–(3) we have, respectively:

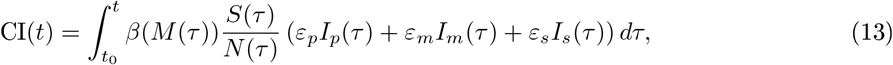

where *β*(*M*) is given in (7), and

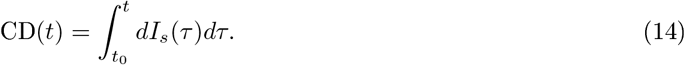

In Fig. 4 the time evolution in [*t*_0_,*t_f_*] of CI(*t*) and CD(*t*) is shown (first and fourth panel from the left), along with that of quarantined individuals *Q*(*t*) (second panel) and hospitalized *I_s_*(*t*) (third panel). The role played by information on the public compliance with mitigation measures is stressed by the comparison with the absolute unresponsive case (*α* = *D* = 0 in (1)–(3)). Corresponding dynamics are labelled by black solid and red dashed lines, respectively. In absence of reactivity to information, the cumulative incidence would have been much less impacted by lockdowns restrictions (11.4 · 10^5^ vs 7.8 · 10^5^ on May 18) and the number of quarantined would have been reduced to those forced by surveillance authorities. As a consequence, the *peak* of hospitalized patients would have been about 77% higher and 10 days time-delayed, with a corresponding increase in cumulative death of more than 100%. For all reported dynamics, the detachment between the responsive and unresponsive case starts to be clearly distinguishable after the first step reduction of 64% in transmission rate (on March 12).

**Figure 4:**
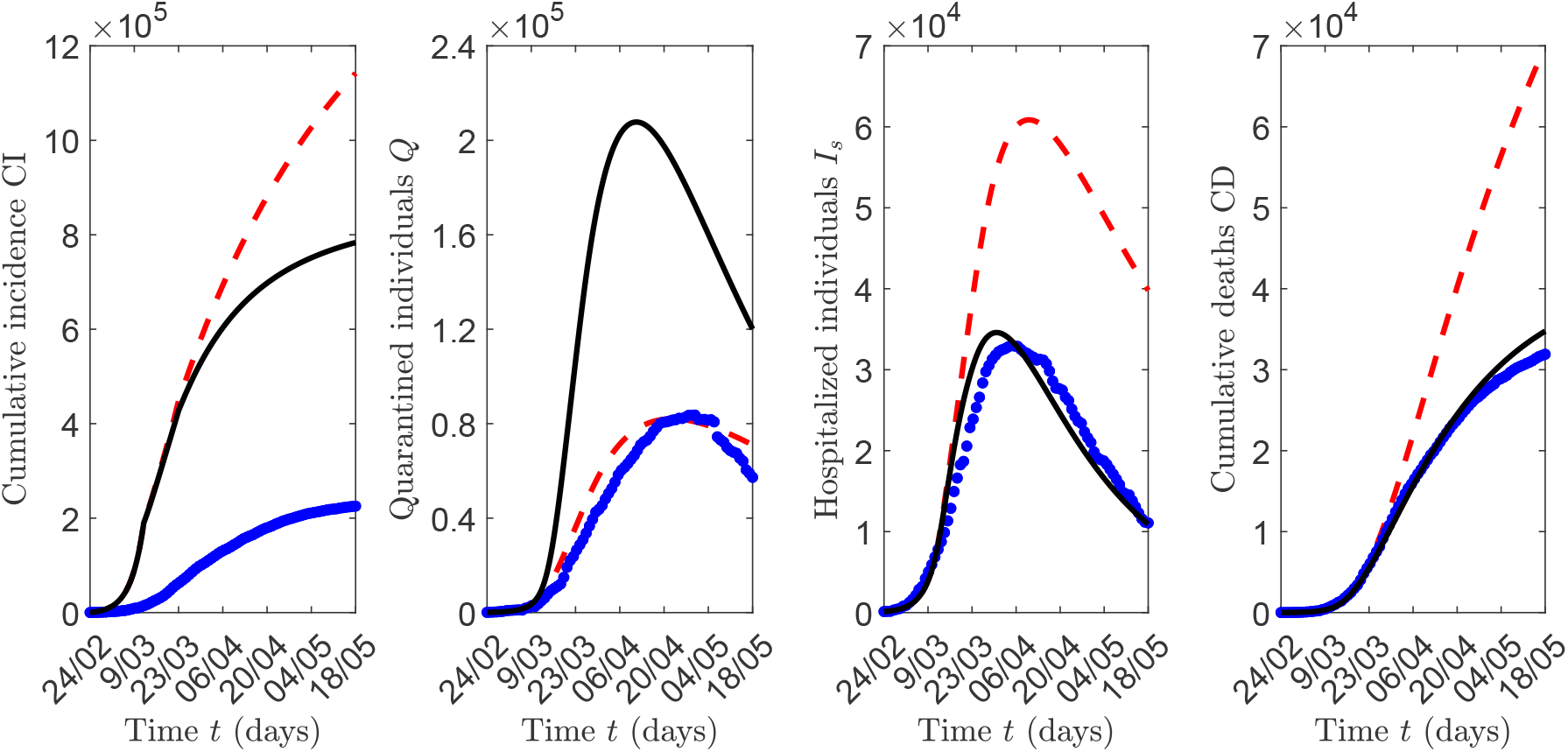
Epidemic evolution predicted by model (1)–(3): Cumulative incidence (first panel from the left), quarantined individuals (second panel), hospitalized individuals (third panel) and cumulative deaths (fourth panel) (black solid lines). The predicted evolution is compared with the case *α* = *D* = 0 (red dashed lines) and with official data (blue dots). Parameter values are given in Tables 1. 2 and 3.

Trends are also compared with officially disseminated data [33] (Fig. 4, blue dots), which seem to conform accordingly for most of the time horizon, except for CI, that suffers from an inevitable and probably high under-estimation [24,25,38,42]. As of May 18, 2020, we estimate about 780,000 contagions, whereas the official count of confirmed infections is 225,886 [33].

We now investigate how the information parameters *k* and *a* may a_ect the epidemic course. More precisely, we assess how changing these parameters affects some relevant quantities: the *peak* of quarantined individuals, max(*Q*) (i.e., the maximum *value* reached by the quarantined curve in [*t*_0_,*t_f_*]), the peak of hospitalized individuals, max(*I_s_*), the cumulative incidence CI(*t_f_*) evaluated at the last day of the considered time frame, i.e. *t_f_* = 84 (corresponding to May 18, 2020), and the cumulative deaths CD(*t_f_*). The results are shown in the contour plots in Fig. 5. As expected, CI(*t_f_*), max(*I_s_*) and CD(*t_f_*) decrease proportionally to the information coverage *k* and inversely to the information delay *a*^−1^: they reach the minimum for *k* = 1 and *a*^−1^ = 1 days. Differently, the quantity max(*Q*) may not monotonically depend on *k* and *a*^−1^ as it happens for *k* ≥ 0.6 and *a*^−1^ ≤ 15 days (see Fig. 5, second panel, lower right corner). In such parameter region, for a given value of *k* (resp. *a*) there are two different values of *a* (resp. *k*) which correspond to the same value of max(*Q*). The absolute maximum 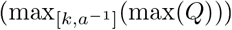 is obtained for *k* = 1 and *a*^−1^ ≈ 7 days. Note that the couple of values *k* = 1, *a*^−1^ = 1 days corresponds to the less severe outbreak, but not with the highest peak of quarantined individuals.

**Figure 5:**
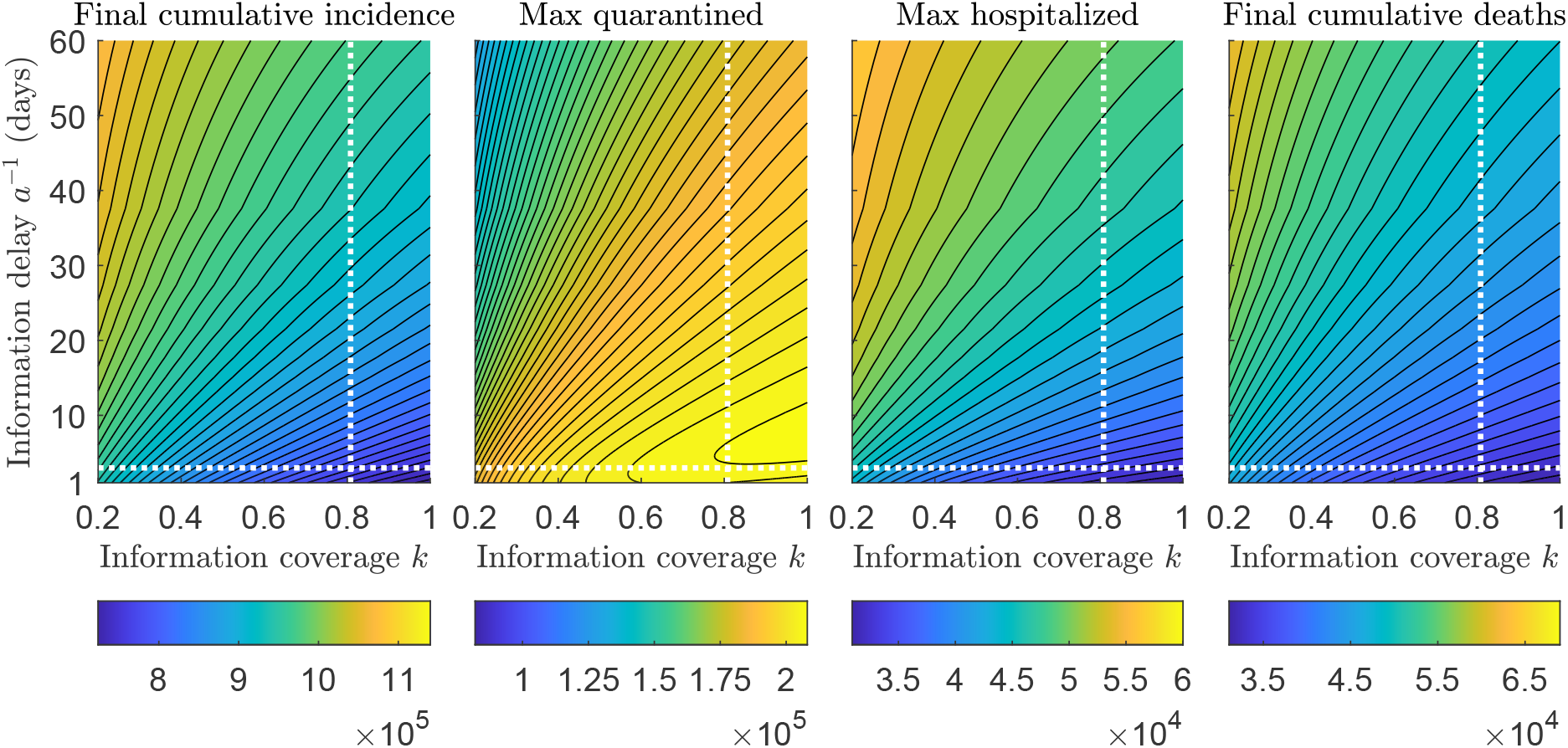
Contour plots of relevant quantities versus information coverage *k* and average delay 1/*a*. First panel from the left: cumulative incidence CI(*t_f_*) evaluated at the last day of the considered time frame, i.e. *t_f_* =84, corresponding to May 18, 2020. Second panel: peak of quarantined individuals. Third panel: peak of hospitalized individuals. Fourth panel: cumulative deaths CD(*t_f_*). The intersection between dotted white lines indicates the values corresponding to the baseline scenario *k* = 0.8, *a* = 1/3 days^−1^. Other parameter values are given in Tables 1, 2 and 3.

In the next, we compare the relative changes for these quantities w.r.t the case when circulating information does not affect disease dynamics. In other words, we introduce the index

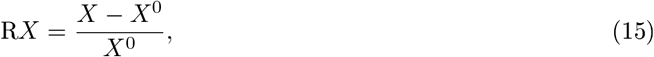

which measures the percentage *relative* change of *X* ∈ {CI(*t_f_*), max(*Q*), max(*I_s_*),CD(*t_f_*)} w.r.t the corresponding quantity *X*^0^ predicted by model (1)–(3) with *α* = *D* = 0.

All the possible values arising in the parameter ranges *k* ∈ [0.2,1] and *a*^−1^ ∈ [1, 60] days are shown in Fig. 6. However, we report in Table 4 three exemplary cases, the baseline and two extremal ones:

**Figure 6:**
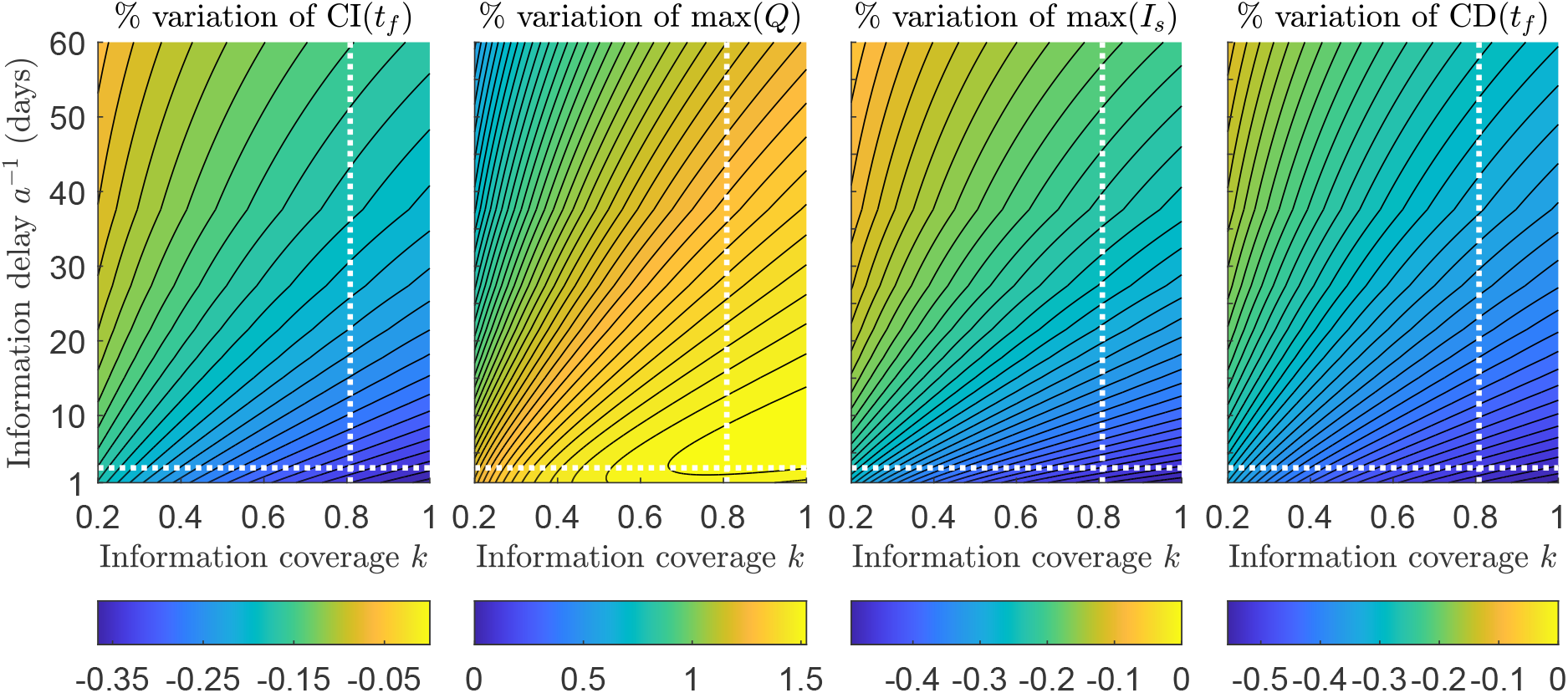
Contour plots of the percentage relative change (15) w.r.t the case *α* = *D* = 0 versus information coverage *k* and average delay 1/*a*. First panel from the left: cumulative incidence CI (*t_f_*) evaluated at the last day of the considered time frame, i.e. *t_f_* = 84, corresponding to May 18, 2020. Second panel: peak of quarantined individuals. Third panel: peak of hospitalized individuals. Fourth panel: cumulative deaths CD(*t_f_*). The intersection between dotted white lines indicates the values corresponding to the baseline scenario *k* = 0.8, *a* = 1/3 days^−1^. Other parameter values are given in Tables 1, 2 and 3.

**Table 4:** Exact and relative values of final cumulative incidence, CI(*t_f_*), quarantined peak, max(*Q*), hospitalized peak, max(*I_s_*), and final cumulative deaths, CD (*t_f_*), for three combinations of information parameters *k* and *a*^−1^ (second to fourth line), in comparison with the information-independent case: *α* = *D* = 0 in (1)–(3) (first line). Other parameters values are given in Tables 1, 2, 3.

| Case | CI($t_f$) | RCI($t_f$) | max($Q$) | Rmax($Q$) | max($I_s$) | Rmax($I_s$) | CD($t_f$) | RCD($t_f$) |
| --- | --- | --- | --- | --- | --- | --- | --- | --- |
| $\alpha = D = 0$ | $11.42 \cdot 10^5$ | 0 | $0.82 \cdot 10^5$ | 0 | $6.09 \cdot 10^4$ | 0 | $6.99 \cdot 10^4$ | 0 |
| $k = 0.8$<br>$a^{-1} = 3$ days | $7.82 \cdot 10^5$ | -0.32 | $2.08 \cdot 10^5$ | 1.53 | $3.45 \cdot 10^4$ | -0.43 | $3.47 \cdot 10^4$ | -0.50 |
| $k = 1$<br>$a^{-1} = 1$ days | $7.24 \cdot 10^5$ | -0.37 | $2.04 \cdot 10^5$ | 1.49 | $3.09 \cdot 10^4$ | -0.49 | $3.11 \cdot 10^4$ | -0.56 |
| $k = 0.2$<br>$a^{-1} = 60$ days | $10.79 \cdot 10^5$ | -0.06 | $1.29 \cdot 10^5$ | 0.57 | $5.70 \cdot 10^4$ | -0.06 | $6.19 \cdot 10^4$ | -0.11 |

i. the baseline scenario *k* = 0.8, *a*^−1^ =3 days, representing a rather accurate and short-delayed communication;
ii. the case of highest information coverage and lowest information delay, *k* = 1, *a*^−1^ = 1 days;
iii. the case of lowest information coverage and highest information delay, *k* = 0.2, *a*^−1^ = 60 days.

Under circumstances of very quick and fully accurate communication (case *(ii)*), Cl (*t_f_*), max(*I_s_*) and CD (*t_f_*) may reduce till 37%, 49% and 56%, respectively (see Table 4, third line). On the other hand, even in case of low coverage and high delay (case *(iii)*), the information still has a not negligible impact on disease dynamics: final cumulative incidence and hospitalized peak reduce till 6%, final cumulative deaths till 11% and quarantined peak increases of 57% about (Table 4, fourth line).

As mentioned above, information and rumors regarding the status of the disease in the community affect the transmission rate *β*(*M*) (as given in (7)) and the quarantine rate γ(*m*) (as given in (8)).

In our last simulation we want to emphasize the role of the information coverage on the quarantine and transmission rates. In Fig. 7 a comparison with the case of low information coverage, *k* = 0.2, is given assuming a fixed information delay *a*^−1^ = 3 days (blue dotted lines). It can be seen that more informed people react and quarantine: an increasing of the maximum quarantine rate from 0.32 to 0.69 days^−1^ (which is also reached a week earlier) can be observed when by increasing the value of *k* till *k* = 1 (Fig. 7, second panel).

**Figure 7:**
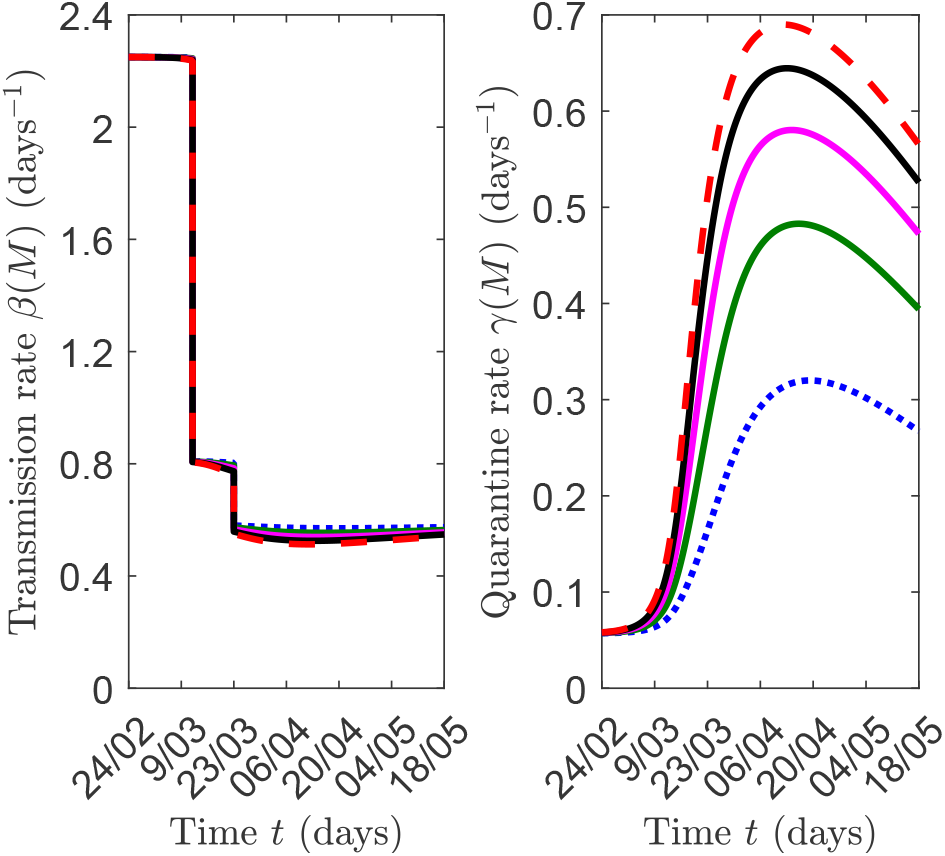
Time evolution of information dependent transmission rate (left panel) and quarantine rate (right panel) of model (1)–(3), by fixing *a*^−1^ = 3 days. Blue dotted lines: *k* = 0.2; green solid lines: *k* = 0.4; magenta solid lines: *k* = 0.6; black solid lines: *k* = 0.8; red dashed lines: *k* =1. Other parameter values are given in Tables 1. 2. 3.

The effect of social behavioral changes is less evident in the transmission rate where increasing the information coverage produces a slight reduction of the transmission rate mainly during the full lockdown phase (Fig. 7, first panel). This reflects the circumstance that the citizens compliance with social distancing is not enhanced by the information-induced behavioral changes during the first stages of the epidemic. On the other hand, a widespread panic reaction may lead people to ‘do it as long as you can’ (see, for example, the case of stormed supermarkets at early stage of the epidemic [48]).

## 6 Conclusions

In this work we propose a mathematical approach to investigate the effects on the COVID–19 epidemic of social behavioral changes in response to lockdowns.

Starting from a SEIR-like model, we assumed that the transmission and quarantine rates are partially determined on voluntary basis and depend on the circulating information and rumors about the disease, modeled by a suitable time-dependent *information index*. We focused on the case of COVID–19 epidemic in Italy and explicitly incorporated the progressively stricter restrictions enacted by Italian government, by considering two step reductions in contact rate (the partial and full lockdowns).

The main results are the followings:

- we estimated two fundamental information-related parameters: the information coverage regarding the daily number of quarantined and hospitalized individuals (i.e. the parameter *k*) and the information delay (the quantity *a*^−1^). The estimate is performed by fitting the model’s solution with official data. We found *k* = 0.8, which means that the public was aware of 80% of real data and *a*^−1^ = 3 days, the time lag necessary to information to reach the public;
- social behavioral changes in response to lockdowns played a decisive role in curbing the epidemic curve: the combined action of voluntary compliance with social distance and quarantine resulted in preventing a duplication of deaths and about 46% more contagions (i.e. approximately 360,000 more infections and 35,000 more deaths compared with the total unresponsive case, as of May 18, 2020);
- even under circumstances of low information coverage and high information delay (*k* = 0.2, *a* = 1/60 days^−1^), there would have been a beneficial impact of social behavioral response on disease containment: as of May 18, cumulative incidence would be reduced of 6% and deaths of 11% about.

Shaping the complex interaction between circulating information, human behavior and epidemic disease is challenging. In this manuscript we give a contribution in this direction. We provide an application of the information index to a specific field case, the COVID 19 epidemic in Italy, where the information dependent model is parametrized and the solutions compared with official data.

Our study presents limitations that leave the possibility of future developments. In particular: *(i)* the model captures the epidemics at a country level but it does not account for regional or local differences and for internal human mobility (the latter having been crucial in Italy at early stage of COVID–19 epidemic); *(ii)* the model does not explicitly account of ICU admissions. The limited number of ICU beds constituted a main issue during the COVID–19 pandemics [22]. This study did not focus on this aspect but ICU admissions could be certainly included in the model; *(iii)* the model could be extended to include age structure. Age has been particularly relevant for COVID–19 lethality rate (in Italy the lethality rate for people aged 80 or over is more than double the average value for the whole population [28]).

Further developments may also concern the investigation of optimal intervention strategies during the COVID–19 epidemics and, to this regard, the assessment of the impact of vaccine arrival. In this case, the approach of information-dependent vaccination could be employed [7,8,18,54].

## A The information index

Consider the scenario of an epidemic outbreak that can be addressed by the public health system through campaigns aimed at raising public awareness regarding the use of protective tools (for example, vaccination, social distancing, bed-net in case of mosquito-borne diseases, etc). Assume also that the protective actions are not mandatory for the individuals (or else, they are mandatory but local authorities are unable to ensure a fully respect of the rules). Then, the final choice to use or not use the protective tools is therefore partially or fully determined by the available information on the state of the disease in the community.

The idea is that such information takes time to reach the population (due to time-consuming procedures such as clinical tests, notification of cases, the collecting and propagation of information and/or rumors, etc) and the population keeps the memory of the past values of the infection (like prevalence or incidence). Therefore, according to the idea of information-dependent epidemic models [40,54], an *information index M* should be considered, which is defined in terms of a delay *τ*, a memory kernel *K* and a function 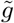 which describes the information that is relevant to the public in determining its final choice to adopt or not to adopt the protective measure (the *message* function).

Therefore, the information index is given by the following distributed delay:

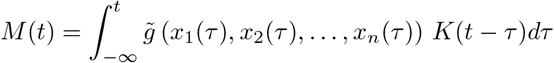

Here, the message function 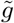 depends generically on the state variables, say *x*_1_ *x*_2_, …, *x_n_* but it may specifically depend only on prevalence [8,15–17], incidence [9] or other relevant quantities like the vaccine side effects [18]. One may assume that:

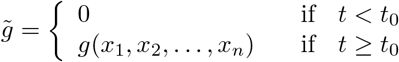

The delay kernel *K*(·) in (2) is a positive function such that 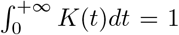. It represents the weight given to past history of the disease. The Erlangian family *Erl_n_,_a_*(*t*) is a good candidate for delay kernel since it may represent both an exponentially fading memory (when *n* =1) and a memory more focused in the past (when *n* > 1). Moreover, when an Erlangian memory kernel is used, one can apply the so called linear chain trick [39] to obtain a system ruled by ordinary differential equations. For example, in the case of exponentially fading memory (or *weak kernel Erl*_1_,*_a_*(*t*)), the dynamics of the information index is ruled by

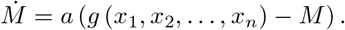

For further details regarding the information index, see [40,54].

## B The *next generation* matrix method

Following the procedure and the notations in [13,51], we prove that the control reproduction number of model (1)–(3), 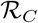, is given by (11). Similarly one can prove that the basic reproduction number is given by (10).

Let us consider the r.h.s. of equations (1b)–(1c)–(1d)–(1e)–(1f), and distinguish the new infections appearance from the other rates of transfer, by defining the vectors

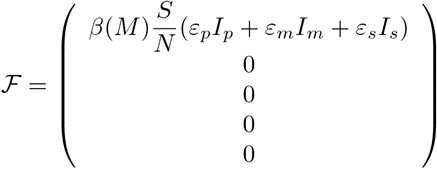

and

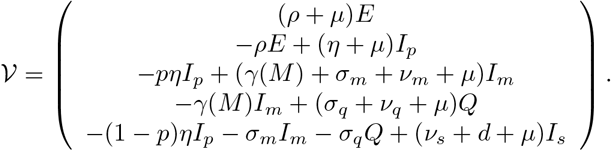

The Jacobian matrices of 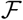 mid 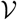 evaluated at model (1)–(3) disease-free equilibrium

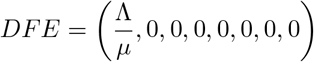

read, respectively,

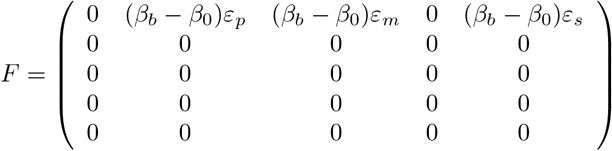

and

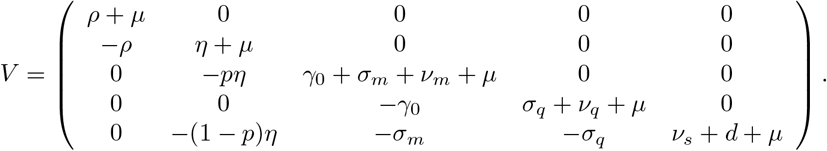

As proved in [13,51], the control reproduction number is given by the spectral radius of the *next generation* matrix *FV*^−1^. It is easy to check that *FV*^−1^ has positive elements on the first row, being the other ones null. Thus, 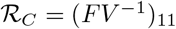, as given in (11).

## Data Availability

there are no external datasets or supplementary material online for this manuscript

## Acknowledgements

The present work has been performed under the auspices of the Italian National Group for the Mathematical Physics (GNFM) of National Institute for Advanced Mathematics (INdAM).

